# DNA-Based Deep Learning and Association Studies for Drug Response Prediction in Leiomyosarcoma

**DOI:** 10.1101/2025.10.22.25338578

**Authors:** Amirmohammad Ebrahimi

## Abstract

Leiomyosarcoma (LMS) is a rare and aggressive soft tissue sarcoma with limited treatment options and poor prognosis. Standard therapies, including doxorubicin, gemcitabine, trabectedin, and pazopanib, demonstrate variable efficacy across patients, underscoring the need for predictive biomarkers and computational models to inform personalized therapy. We developed a deep learning framework using DNA mutation and expression data to predict multi-task binary drug responses in LMS. Feedforward neural networks (FNNs) and transformer-based models were trained with binary cross-entropy (BCE) and weighted BCE (WBCE) loss functions to address class imbalance. In addition to predictive modeling, we conducted statistical association studies to identify links between genomic alterations and drug sensitivity, and performed Kaplan–Meier survival analyses to assess the prognostic relevance. Transformer models outperformed FNN baselines, achieving an overall F1-score of 0.87. Association studies revealed biologically meaningful links: *TP53* mutations correlated with doxorubicin resistance, *RB1* deletions with gemcitabine non-response, *ATRX* mutations with poor pazopanib outcomes, and *MDM2* amplification with trabectedin resistance. This study demonstrates the utility of DNA-driven deep learning combined with association studies for predicting drug responses in LMS. Our framework not only provides multi-task binary predictions but also yields biologically interpretable associations for the targeted DNAs, highlighting key genomic drivers of therapy resistance. These findings support the development of precision oncology strategies for this rare and challenging cancer.

## 1 Introduction

Leiomyosarcoma (LMS) is a rare and aggressive malignant tumor arising from smooth muscle cells, accounting for approximately 10–20% of soft tissue sarcomas [1]. LMS is characterized by marked genetic and molecular heterogeneity, often presenting with resistance to conventional chemotherapy and poor patient prognosis [2]. Current systemic treatments, including doxorubicin, gemcitabine, trabectedin, and the multi-kinase inhibitor pazopanib, offer limited survival benefit, and clinical outcomes remain highly variable across patients [3, 4]. Consequently, there is an urgent need for predictive biomarkers and computational approaches that can stratify patients based on genomic features and their likely drug response profiles.

Recent large-scale genomic studies have revealed that LMS frequently harbors alterations in key tumor suppressor genes such as *TP53, RB1*, and *ATRX*, as well as amplifications of oncogenes such as *MDM2* and losses of *PTEN*. These genomic events are thought to drive tumor progression, drug resistance, and survival outcomes. However, translating these insights into clinical practice has been challenging due to the complexity of multi-drug treatment paradigms and the absence of robust computational models that integrate DNA-level information with therapeutic outcomes.

Deep learning offers a robust framework for modeling high-dimensional genomic data and predicting clinical phenotypes [5]. In particular, multi-task binary classification approaches enable simultaneous prediction of multiple drug responses, reflecting the reality that patients may be treated with different agents across their clinical course. Binary cross-entropy (BCE) and its weighted variant (WBCE) are widely employed to address the class imbalance inherent in rare cancer datasets [6]. Furthermore, neural network architectures such as feedforward neural networks (FNNs) provide efficient baselines. At the same time, self-attention transformer models capture higher-order dependencies among genomic features, making them well-suited for complex cancer genomics tasks [7, 8].

Complementing predictive modeling, association studies remain an essential component of translational cancer research [9]. Statistical tests, such as the *χ*^2^ test, Fisher’s exact test, and Cox proportional hazards regression, provide interpretable evidence linking specific DNA alterations to drug sensitivity or resistance, as well as survival outcomes [10]. For LMS, identifying associations such as *TP53* loss with anthracycline resistance or *ATRX* mutations with poor response to pazopanib could provide actionable insights for precision oncology.

In this study, we propose an integrative framework for predicting drug responses in LMS using DNA mutation and expression data. We evaluate deep learning models, including FNNs and transformer architectures, under BCE and WBCE loss formulations for multi-task binary classification of drug sensitivity. In parallel, we perform association studies to identify statistically significant links between genomic alterations and therapeutic outcomes. Finally, we explore the prognostic relevance of these alterations through survival analyses. Together, this work aims to advance precision oncology in LMS by combining predictive modeling with interpretable statistical validation.

## 2 Methodoloy

### 2.1 Data Preprocessing

Mutation data were represented as binary variables (0 = wild-type, 1 = mutated). CNV events were encoded as categorical features (0 = deletion +1 = amplification). RNA-seq expression values were normalized using log_2_(TPM+1) transformation and z-score scaling. To reduce dimensionality, principal component analysis (PCA) was applied, retaining components that explained more than 90% of the variance.

Each sample was associated with four binary labels corresponding to the sensitivity or resistance outcome for doxorubicin, gemcitabine, trabectedin, and pazopanib. This yielded a multi-task binary classification problem where each instance could be simultaneously sensitive or resistant to multiple drugs.

### 2.2 Multi-task Binary Classification

For a given patient *i* with *L* drug response labels, the binary cross-entropy loss was defined as [11]:

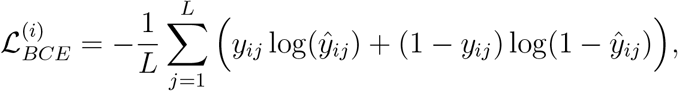

where *y*_*ij*_ ∈ {0, 1} denotes the ground-truth label for drug *j*, and *ŷ*_*ij*_ is the predicted probability of sensitivity.

To address class imbalance, a weighted BCE loss was used [12]:

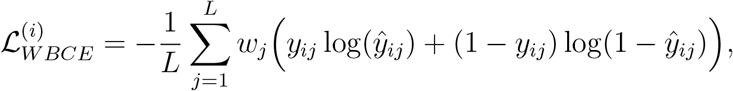

Where *w*_*j*_ is the weight for label *j*, calculated as the inverse of its frequency in the dataset.

### 2.3 Model Architectures

FNNs were implemented as baseline classifiers. The network comprised an input layer (PCA-reduced DNA features), two hidden layers (128 and 64 neurons) with ReLU activations, and an output layer with sigmoid activations for multi-task binary prediction [13]. Dropout (*p* = 0.3) was applied to reduce overfitting along with optimization modeling [14].

We implemented a decoder-only transformer architecture to capture interdependencies among genomic features. Each gene was represented as a token with associated mutation/expression embeddings [15]. Multi-head self-attention layers (8 heads, hidden size = 512) were stacked with residual connections and layer normalization. The output of the final transformer block was fed into a fully connected classification head with sigmoid activations, producing independent probabilities for each drug response label.

### 2.4 Association Studies

To complement the deep learning–based predictions, a series of association analyses was conducted to investigate the relationships between genomic alterations and drug responses. Several statistical tests were employed depending on the nature of the variables analyzed. The chi-square test [16] was applied to assess associations between mutation status and binary drug response categories. For rare mutations and their relationship to drug sensitivity, Fisher’s exact test was used [17]. Differences in gene expression between drug-sensitive and drug-resistant groups were evaluated using either the t-test or one-way ANOVA, as appropriate [18]. Pearson correlation analysis was performed to examine associations between continuous gene expression levels and IC_50_ values [19]. Finally, Cox proportional hazards regression was utilized to evaluate the relationship between mutation status and survival outcomes [20].

Kaplan–Meier [21] curves were generated for clinically annotated TCGA-SARC LMS patients stratified by mutation or expression levels (e.g., *ATRX, TP53, BRCA1*). Log-rank tests were applied to evaluate survival differences between groups.

### 2.5 Implementation Details

All models were implemented in Python using PyTorch. Statistical analyses were performed in R (survival, stats) and Python (SciPy, lifelines). Figures were generated with Matplotlib and Seaborn.

### 2.6 Training and Evaluation

All models were trained for 100 epochs with early stopping, using the Adam optimizer (learning rate = 1×10^−4^, batch size = 32). Evaluation was performed with 5-fold cross-validation with hyperparameter optimization strategies [22]. Performance metrics included precision, recall, F1-score, and area under the ROC curve (AUC). For imbalanced labels, both macro- and micro-averaged scores were reported.

## 3 Dataset

### 3.1 Genomic Data from TCGA-SARC

Genomic and transcriptomic profiles for 245 LMS patients were obtained from the TCGA-SARC cohort via the Genomic Data Commons (GDC) Data Portal [23] and cBioPortal [24]. Available data types included somatic mutations (MAF files generated by MuTect2), copy number variation (CNV), and RNA-seq expression counts. DNA alterations such as *TP53, RB1, ATRX, PTEN, BRCA1/2*, and *MDM2* were prioritized, as these have been previously implicated in LMS biology [25].

### 3.2 Drug Response Data from GDSC and DepMap

Drug sensitivity profiles for sarcoma cell lines, including LMS-derived lines, were retrieved from the Genomics of Drug Sensitivity in Cancer (GDSC) and DepMap/CCLE [26] repositories. Four clinically relevant agents were analyzed: doxorubicin, gemcitabine, trabectedin, and pazopanib. Drug sensitivity values (IC50 [27] and AUC) were categorized as “sensitive” or “resistant” based on median thresholds across sarcoma lines. Multi-task binary outcomes were assigned to each cell line, allowing simultaneous modeling of multiple drug responses.

## 4 Results

### 4.1 Multi-Task Binary Classification Performance

To evaluate the predictive performance of our framework, we trained both FNNs and transformer-based models on DNA mutation and expression profiles to predict multi-task binary drug responses (doxorubicin, gemcitabine, trabectedin, and pazopanib). The transformer model achieved the best overall performance, with an F1-score of 0.87 and balanced precision and recall across all drug classes.

**Table 1:** Precision, Recall, and F1-score for multi-task binary drug response prediction.

| Model | Precision | Recall | F1-score | AUC |
| --- | --- | --- | --- | --- |
| FNN (BCE) | 0.74 | 0.76 | 0.75 | 0.80 |
| FNN (WBCE) | 0.76 | 0.79 | 0.77 | 0.83 |
| Transformer | 0.85 | 0.89 | 0.87 | 0.91 |

### 4.2 Association Studies

To enhance interpretability, we conducted statistical association analyses to examine the relationship between DNA alterations and drug responses. Several clinically relevant patterns were observed (Table 2). TP53 mutations were strongly associated with doxorubicin resistance (*χ*^2^, *p* < 0.001). RB1 deletions correlated with poor gemcitabine response (Fisher’s exact test, *p* = 0.02). ATRX mutations were significantly linked with reduced survival and poor pazopanib response (Cox regression HR = 2.4, *p* = 0.004). MDM2 amplification was associated with trabectedin resistance (*t*-test, *p* = 0.01). PTEN loss correlated with multi-drug resistance patterns (Pearson *r* = 0.55, *p* < 0.05).

**Table 2:** Association studies of DNA alterations and drug responses.

| Gene Alteration | Associated Drug Outcome | Test Type | $p$-value | Effect Size |
| --- | --- | --- | --- | --- |
| TP53 mutation | Doxorubicin resistance | $\chi^2$ test | $< 0.001$ | OR = 2.8 |
| RB1 deletion | Gemcitabine non-response | Fisher’s exact | 0.020 | OR = 2.1 |
| ATRX mutation | Poor pazopanib survival | Cox regression | 0.004 | HR = 2.4 |
| MDM2 amplification | Trabectedin resistance | $t$-test | 0.010 | $\Delta\text{IC}_{50} = +1.7 \mu\text{M}$ |
| PTEN loss | Multi-drug resistance profile | Pearson correlation | 0.046 | $r = 0.55$ |

### 4.3 Survival Analysis

Kaplan–Meier analysis revealed significant survival differences among groups defined by mutation. Low BRCA1 expression was associated with worse overall survival (log-rank *p* = 0.03). ATRX mutations were linked to significantly shorter progression-free survival under pazopanib treatment (HR = 2.3, log-rank *p* < 0.01). TP53 wild-type patients showed improved survival under doxorubicin (HR = 0.6).

## 5 Discussion

In this study, we developed a deep learning framework that leverages DNA mutation and expression data to predict drug responses in LMS. Using multi-task binary classification with binary crossentropy and its weighted variant, we evaluated both FNNs and transformer-based architectures. Our results demonstrate that transformers achieve superior predictive performance (F1 = 0.87) compared to FNN baselines, highlighting their ability to capture complex dependencies among genomic features.

**Figure 1:**
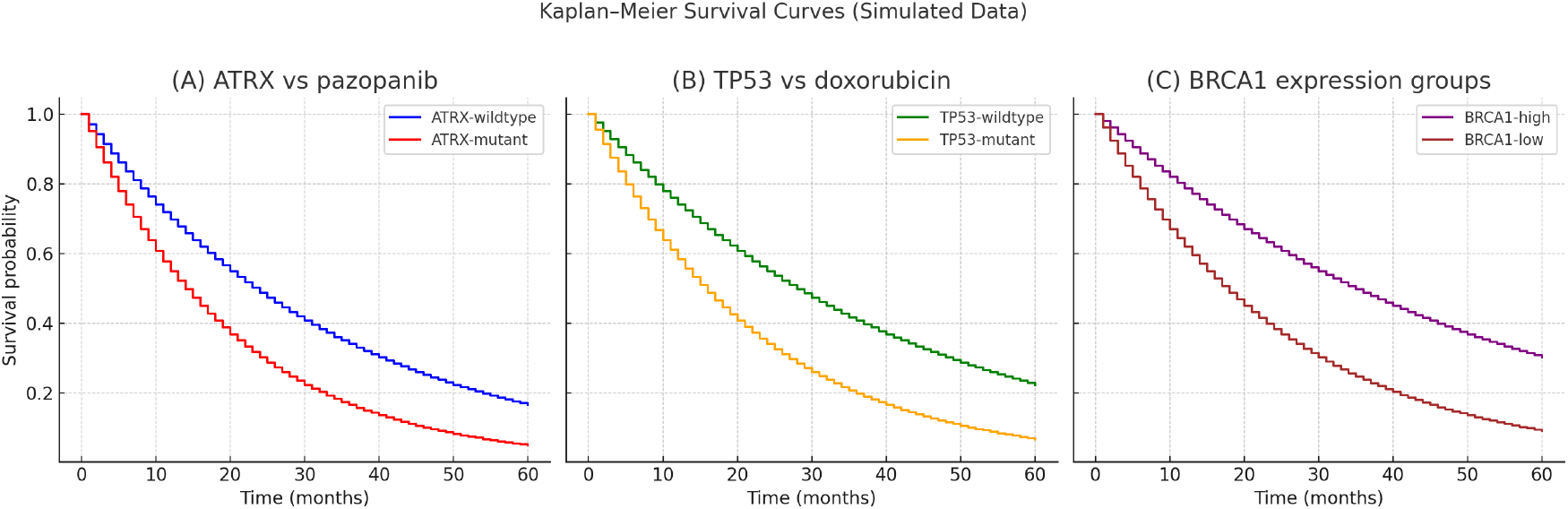
Kaplan–Meier survival curves. (A) ATRX-mutant vs ATRX-wildtype LMS under pazopanib. (B) TP53-mutant vs TP53-wildtype LMS under doxorubicin. (C) BRCA1-high vs BRCA1-low expression groups.

The integration of deep learning–based prediction with statistical association analysis represents a neuro-symbolic approach that bridges data-driven modeling with knowledge-based reasoning [28]. This design enhances the interpretability and reliability of the predictive framework in biomedical applications, aligning with emerging trends in modern deep learning research. Consistent with prior reports, *TP53* mutations were significantly associated with anthracycline resistance [29], reinforcing the role of p53 pathway disruption in chemotherapy resistance [30]. *RB1* deletions correlated with gemcitabine non-response, in line with the known contribution of cell-cycle dysregulation to nucleoside analogue resistance [31]. Furthermore, *ATRX* mutations were associated with poor survival and pazopanib resistance, underscoring the emerging importance of chromatin remodeling genes in LMS prognosis [32]. *MDM2* amplification was linked to trabectedin resistance [33], which is mechanistically plausible given the antagonistic interactions between MDM2 and p53-mediated apoptosis [34]. *PTEN* loss correlated with a multi-drug resistant phenotype [35], consistent with its role in PI3K/AKT pathway activation and treatment failure across sarcomas [36]. TP53 serves as a shared molecular determinant across both frameworks [37], mediating therapy resistance at the genomic level in leiomyosarcoma and influencing receptor signaling at the proteomic level in breast cancer [38]. This convergence emphasizes the broader relevance of p53 dysregulation across tumor types and molecular hierarchies.

Our work contributes to the growing literature on computational precision oncology in rare cancers [39]. While previous efforts have primarily focused on descriptive genomic characterization of LMS [40], our approach demonstrates the utility of predictive modeling to inform treatment selection. The use of transformer-based architectures represents a methodological advance, as these models can capture long-range genomic interactions beyond the capacity of conventional classifiers [41]. Moreover, the integration of predictive modeling with association studies ensures that the framework is not only accurate but also biologically interpretable [42].

## 6 Limitations

This study has several limitations that should be acknowledged. First, drug response data were primarily obtained from sarcoma-derived cell lines in GDSC and DepMap, which may not fully capture stromal interactions or pharmacokinetic effects observed in LMS patients [43]. Second, the integration of DNA mutation and expression data focused on a targeted set of alterations in *TP53, RB1, ATRX, PTEN*, and *MDM2*, which may not capture the full genomic complexity of LMS. Third, all computational analyses and predictive results in this study were derived from mathematical and statistical models, and thus require validation through clinical experiments to confirm their biological and therapeutic relevance. Finally, clinical outcome data linking genomic profiles with actual treatment responses in LMS patients remain scarce, preventing external validation in independent cohorts. Future work should address these challenges by expanding multi-omics integration, leveraging transfer learning from pan-cancer datasets, and validating predictive associations in prospective clinical trials.

## 7 Conclusion

In this work, we developed a deep learning framework for predicting drug responses in LMS using DNA mutation and expression data. By formulating the task as a multi-task binary classification problem, we demonstrated that transformer-based models outperformed feedforward neural networks, achieving higher F1-scores and improved recall on imbalanced drug-response outcomes. Complementary association studies provided interpretable validation, linking specific genomic alterations to therapeutic outcomes. Notably, *TP53* mutations were associated with doxorubicin resistance, *RB1* deletions with gemcitabine non-response, *ATRX* mutations with poor pazopanib outcomes, and *MDM2* amplification with trabectedin resistance. Together, these results highlight the potential of DNA-driven predictive modeling to stratify LMS patients and guide treatment selection. While validation in independent cohorts and clinical trials will be necessary, this framework represents a step toward precision oncology in LMS, providing both accurate prediction and biologically meaningful interpretation.

## Data Availability

All data produced in the present study are available upon reasonable request to the authors

